# Antibodies elicited by the 2025-2026 influenza vaccine in humans

**DOI:** 10.64898/2026.01.05.26343449

**Authors:** Jiaojiao Liu, Shuk Hang Li, Naiqing Ye, Tachianna Griffiths, Elizabeth M Drapeau, Reilly K. Atkinson, Ronald G. Collman, Scott E. Hensley

## Abstract

A new H3N2 variant (named subclade K) possesses several key hemagglutinin substitutions and is circulating widely during the 2025-2026 influenza season. In this report, we completed experiments to determine if the 2025-2026 seasonal influenza vaccine elicits antibodies in humans that recognize this variant. We find that H3N2 subclade K viruses are antigenically advanced; however, the 2025-2026 seasonal influenza vaccine elicited antibodies in many individuals that efficiently recognized these viruses. Thus, the current seasonal influenza vaccine will likely be somewhat effective at preventing H3N2 subclade K virus infections.

## Introduction

Seasonal influenza viruses continuously undergo antigenic drift and vaccine antigens are updated regularly. A new H3N2 variant (named subclade K) emerged in the spring and summer of 2025 and is now circulating widely in the United States and other parts of the world^1^. The hemagglutinin (HA) protein of H3N2 subclade K viruses possesses eleven substitutions (K2N, T135K, S144N, N145S, N158D, I160K, Q173R, A186D, K189R, T328A, HA2: S48N) relative to the HA of the Northern Hemisphere 2025-2026 H3N2 vaccine strain, many of which are in antibody-binding epitopes (Fig. 1A). In September, the World Health Organization reported that ferret anti-sera raised against the current H3N2 vaccine strain poorly react to subclade K H3N2 viruses^2^, raising the concern that there could potentially be a major vaccine mismatch during the 2025-2026 Northern Hemisphere influenza season. H3N2 subclade K viruses circulated widely in England during the fall of 2025, and early season vaccine effectiveness estimates against influenza-related emergency department attendances and hospital admissions were unexpectedly high^3^. It is therefore important to determine if the 2025-2026 influenza vaccine elicits antibodies in humans that recognize H3N2 subclade K viruses.

**Figure 1:**
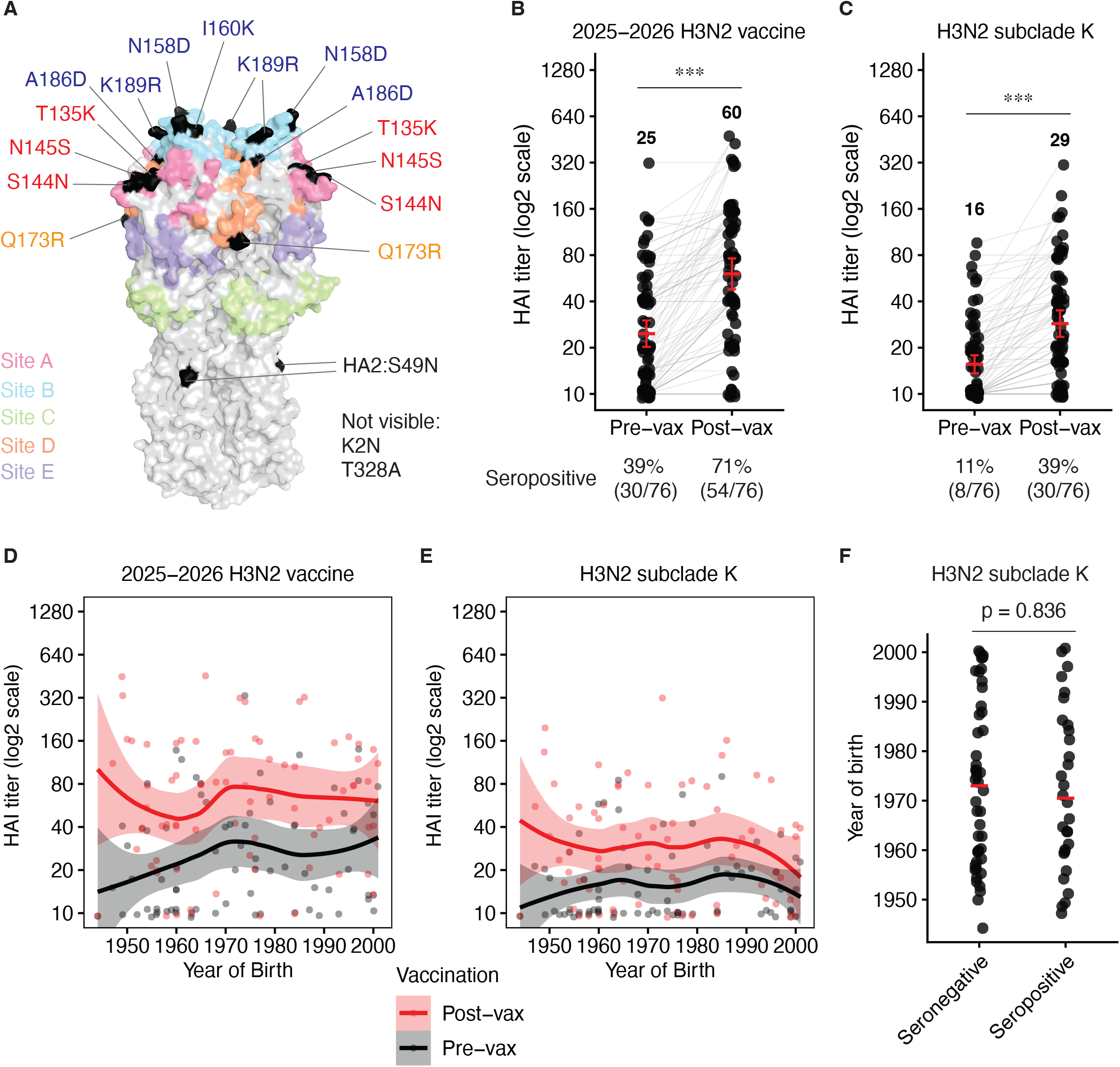
Human antibody recognition of H3N2 subclade K virus. Panel A shows the crystal structure of the A/Victoria/22/2020 HA trimer (PDB_8FAQ) with major antigenic sites (Sites A-E) displayed in different colors. Amino acid differences between the HA of the 2025-2026 H3N2 vaccine strain (A/Croatia/10136RV/2023), and H3N2 subclade K virus (A/New York/GKISBBBG87773/2025) are shown in black. Panels B and C show hemagglutination inhibition (HAI) titers to the 2025-2026 H3N2 vaccine strain (A/Croatia/10136RV/2023; a subclade J.2 virus) and an H3N2 subclade K variant virus (A/New York/GKISBBBG87773/2025) using sera collected from 76 adults before and after (27-30 days) receiving the 2025-2026 influenza vaccine. Each data point in panels B and C represents the geometric mean titer (GMT) using serum from one individual tested in 2 independent experiments. Lines in panels B and C connect data points from the same individual. Seropositivity was defined as an HAI titer ≥40. HAI titers are plotted as the GMT with 95% confidence interval (CI). The Wilcoxon Rank-Sum test was performed to compare HAI titers before vaccination and after vaccination. ^***^ P < 0.001. The same data from panels B and C are displayed in panels D and E with each participant’s birth year indicated on the x axes. The trend lines in panels D and E are locally estimated scatterplot smoothing curves (smoothing parameter = 0.7) with 95% CIs. Panel F shows the birth year distributions among H3N2 subclade K seronegative and seropositive individuals after vaccination. The median year of birth is shown as a red line. The Wilcoxon Rank-Sum test was performed to compare the difference in birth years between seronegative participants and seropositive participants.

## Methods

### Human serum samples

Serum samples from 76 adults (ages 24-81) were collected before and 27-30 days after receiving a standard dose of the egg-based 2025-2026 Flulaval Trivalent influenza vaccine (GaxoSmithKline) between October-November 2025. This study was approved by the Institutional Review Board of the University of Pennsylvania under protocol #849398.

### Viruses

Influenza viruses were generated as previously described^4^. Nucleic acids for the HA and NA of the 2025-2026 H3N2 vaccine strain (A/Croatia/10136RV/2023; GISAID accession numbers EPI_ISL_19296516) and an H3N2 subclade K virus (A/New York/GKISBBBG87773/2025; GISAID accession numbers EPI_ISL_20126669) were synthesized and cloned into the PHW2000 reverse genetics plasmid (Twist Bioscience). Viruses were rescued by transient transfection of the HA, NA, and 6 A/Puerto Rico/8/1934 internal genes (which were cloned into PHW2000 reverse genetics plasmids) into co-cultures of 293T and MDCK-SIAT1 cells. The 2025-2026 H3N2 vaccine strain (A/Croatia/10136RV/2023) was then propagated in 10-day-old fertilized chicken eggs, and the H3N2 subclade K virus (A/New York/GKISBBBG87773/2025) was passaged twice and propagated in MDCK-SIAT1 cells. All virus stocks were stored at -80 °C, and the sequences were confirmed after propagation.

### HAI assays

HAI assays were performed as previously described^5^. HAI titrations were performed in round-bottom 96-well plates. Serum samples were treated with receptor-destroying enzyme (RDE) (Denka-Seiken) at 37°C for 2 hours and heat-inactivated at 56°C for 30 minutes. The samples were then incubated with a 1.1 volume 10% (v/v) turkey red blood cell solution at 4°C for 1 hour. Sera were 2-fold serially diluted in DPBS with a 1:20 starting dilution, and four agglutinating doses of viruses were added in a total volume of 100 μl. Then, 12.5 μl of a 2% (v/v) turkey red blood cell solution was added to each sample. After 1 hour of incubation at room temperature, agglutination was determined. HAI titers were recorded as the inverse of the highest dilution that inhibited hemagglutinin of turkey red blood cells. Serum samples with a titer of less than 1:20 were assigned an HAI titer of 1:10. We completed two independent replicates with each sera sample.

### Statistical analyses

The Wilcoxon Rank-Sum test was used to compare HAI titers before and after vaccination, as well as to examine the difference in year of birth between seronegative and seropositive participants. The trend lines of HAI titers by year of birth are locally estimated scatterplot smoothing curves (smoothing parameter = 0.7, degree = 1) with 95% CIs.

## Results

We collected sera from 76 individuals before and ∼1 month (between 27-30 days) after receiving a standard dose of the egg-based 2025-2026 Flulaval Trivalent influenza vaccine. We enrolled participants with diverse birth years, since early childhood influenza exposures can affect the specificity of antibodies elicited by contemporary influenza vaccine strains^6,7^. We completed hemagglutination inhibition (HAI) assays to identify antibodies that block virus attachment of the 2025-2026 H3N2 vaccine strain (A/Croatia/10136RV/2023; a subclade J.2 virus) and an H3N2 subclade K variant virus (A/New York/GKISBBBG87773/2025). Antibody titers against both A/Croatia/10136RV/2023 and A/New York/GKISBBBG87773/2025 increased in sera from most individuals following vaccination; however, antibody geometric mean titers were approximately 2-fold higher to A/Croatia/10136RV/2023 compared to A/New York/GKISBBBG87773/2025 after vaccination (Fig. 1B-C). Before vaccination, 39% (30 of 76) of participants were seropositive (≥40 HAI titer) against A/Croatia/10136RV/2023 and only 11% (8 of 76) of participants were seropositive against A/New York/GKISBBBG87773/2025. Following vaccination, 71% (54 of 76) and 39% (30 of 76) of participants were seropositive against A/Croatia/10136RV/2023 and A/New York/GKISBBBG87773/2025 respectively. We did not observe major birth year-related differences in antibody reactivity to either virus (Fig. 1D-E), and we observed similar birth year distributions among H3N2 subclade K seronegative and seropositive individuals following vaccination (Fig. 1F).

## Conclusions

Collectively, our data suggest that H3N2 subclade K viruses are antigenically advanced compared to the 2025-2026 H3N2 vaccine strain; however, the antigenic differences that we observed in sera from some humans are not as large as previously reported in ferrets^2^. While we find that human antibodies elicited by vaccination react more efficiently to the H3N2 vaccine strain relative to subclade K H3N2 viruses, we found that many individuals produced antibodies that efficiently recognized subclade K H3N2 viruses after vaccination. Our study highlights the benefits of receiving influenza vaccinations, even in seasons that include circulation of variant viruses.

## Data Availability

All data are provided in this manuscript.

